# The impact of universal glove and gown use on *Clostridioides difficile* acquisition, a cluster randomized trial

**DOI:** 10.1101/2022.04.11.22273721

**Authors:** Daniel J. Morgan, Erik R. Dubberke, Tiffany Hink, Gwen Paszkiewicz, Carey-Ann D. Burnham, Lisa Pineles, Larry Magder, J. Kristie Johnson, Surbhi Leekha, Anthony D. Harris

**Author notes:** **Corresponding author:** Daniel J. Morgan, MD MS, 10 S. Pine St., MSTF 334, Baltimore, MD 21201. **Trial registration:** Clinicaltrials.gov identifier NCT01318213.

## Abstract

**Importance:** *Clostridioides difficile* is the most common cause of healthcare-associated infections (HAI) in the US. It is unknown whether universal gown and glove use in intensive care units (ICUs) decreases acquisition of *C. difficile*.

**Objective:** To assess whether wearing gloves and gowns for all patient contact in the ICU decreases acquisition of *C. difficile* compared with usual care.

**Design, setting, and Participants:** Secondary analysis of a cluster-randomized trial in 20 medical and surgical ICUs in 20 US hospitals from January 4, 2012, to October 4, 2012.

**Interventions:** After a baseline period, ICUs were randomized to standard practice for glove and gown use vs. the intervention of all healthcare workers being required to wear gloves and gowns for all patient contact and when entering any patient room (contact precautions).

**Main outcomes and measures:** The primary outcome was acquisition of toxigenic *C. difficile* determined by surveillance cultures collected on admission and discharge from the ICU. Secondary outcomes included ribotype 027-like *C. difficile* acquisition and the impact of other factors on acquisition.

**Results:** From the 26,749 patients enrolled in the study, a total of 21,845 patients had both admission and discharge perianal swabs cultured for toxigenic *C. difficile*. On admission, 9.43% (2,060/21,845) of patients were colonized with toxigenic *C. difficile*. No significant difference was observed in the rate of toxigenic *C. difficile* acquisition with universal gown and glove use. Differences in acquisition rates in the study period compared to baseline period in control ICUs were 1.49 per 100 patient days vs 1.68 per 100 patient days in universal gown and glove ICUs, (rate difference -0.28, generalized linear mixed model, p=0.091). Similarly, there was no difference in rates of ribotype 027-like *C. difficile* acquisition: control ICUs 0.13 per 100 patient days vs. 0.16 per 100 patient days in universal gown and glove ICUs during the study period, (rate difference -0.03, generalized linear mixed model, p=0.35). Secondary analyses identified *C. difficile* colonization was associated with acquisition, p=0.014).

**Conclusions and relevance:** Glove and gown use for all patient contact in medical and surgical ICUs did not result in a reduction in the acquisition of *C. difficile* compared with usual care.

**Key Points:** *Question:* Does wearing gloves and gowns for all patient contact in the ICU decrease acquisition of *C. difficile?*

*Findings:* Glove and gown use for all patient contact in medical and surgical ICUs did not result in a difference in the acquisition of *C. difficile* compared with usual care.

*Meaning:* Glove and gown use does not impact acquisition of *C. difficile* from asymptomatically colonized patients. Universal use of glove and gowns or targeted use based on active surveillance for *C. difficile* are unlikely to impact transmission.

## BACKGROUND

*Clostridioides difficile* (*C. difficile)* is the most common cause of healthcare-associated infections (HAIs) in the US, causing at least 450,000 infections and 29,000 associated deaths per year.^1^ Current *C. difficile* infection (CDI) prevention strategies focus on preventing *C. difficile* transmission only from patients with symptomatic CDI and ignore the much more common asymptomatic *C. difficile* carriers.^2,3^ Asymptomatic *C. difficile* carriers have been shown to contaminate the hospital environment, transfer *C. difficile* spores to healthcare personnel hands, and introduce strains associated with subsequent CDI cases.^1^ In addition, recent studies suggest at least 30% to 50% of new CDI cases may be the result of transmission from asymptomatic *C. difficile* carriers.^4–6^ However, the potential impact of interventions directed at asymptomatic *C. difficile* carriers has not been adequately studied. One such strategy to prevent *C. difficile* transmission among hospitalized patients would be the use of gloves and gowns (contact precautions) for patients asymptomatically colonized with *C. difficile*. Identifying reductions in *C. difficile* transmission from asymptomatic carriers with contact precautions would change the paradigm of *C. difficile* prevention. Small observational studies have explored this approach with active surveillance testing at admission and found associations with lower rates of CDI.^7,8^ However, no randomized trials have assessed the impact of contact precautions on *C. difficile* acquisition.

Controversy exists about the relative advantages and disadvantages of contact precautions.^9,10^ Previously, we published a cluster-randomized trial titled the Benefits of Universal Glove and Gowning Study (BUGG) that showed that wearing gowns and gloves for all patients resulted in a reduction in Methicillin-resistant *Staphylococcus aureus* (MRSA) acquisition, but no effect on the composite primary outcome of MRSA or Vancomycin resistant *Enterococcus* (VRE) acquisition.^11^ The study also showed no increase in adverse events and improved hand hygiene compliance on room exit with the intervention. Likewise, no statistically significant impact was seen on acquisition of gram-negative pathogens.^12^

In the current study, we used previously collected and stored perianal samples from the BUGG cluster-randomized trial to assess if wearing gloves and gowns for all patient contact in the ICU reduces acquisition rates of *C. difficile*.

## METHODS

### Study design

This study is a secondary analysis of specimens collected in the BUGG study, a 20-hospital cluster-randomized trial of universal glove and gown compared to standard practice. The study was conducted in medical, surgical and medical-surgical ICUs varying in size from nine to 36 beds and located across the United States in rural, urban, academic and non-academic settings. The primary outcome of the original trial was acquisition of MRSA or VRE. Details of the original study design have been previously published.^11^ The study had a baseline period from September 1, 2011 to December 31, 2011. After the baseline period, ICUs were randomized to either the intervention or control arm. The study period was from January 4, 2012 to October 4, 2012. The trial was conducted in accordance with the Consolidated Standards of Reporting Trials (CONSORT) guidelines (Supplemental Figure 1).^13^

A total of 45,821 peri-anal swabs were obtained in the BUGG study. These peri-anal cultures were collected on admission and discharge for all eligible participants and shipped to the University of Maryland laboratory. We previously published this microbiology shipping and processing methodology, demonstrating excellent recovery of bacteria.^14,15^ All peri-anal swabs were frozen in tryptic soy broth containing 15% glycerol at -80°C to allow for future recovery of organisms. Recovery of organisms from frozen swabs has been shown by our group and others to be between 91% and 98%. We cultured each frozen swab for *C. difficile* in anaerobic conditions. All *C. difficile* isolates were then frozen and shipped to Washington University for toxin and *tcdC* gene characterization. Isolates were subcultured on pre-reduced blood agar and identification was confirmed with Matrix-Assisted Laser Desorption/Ionization-Time Of Flight (MALDI-TOF) (VITEK MS; bioMérieux, Durham, NC, USA). The presence of *tcdA, tcdB, cdtA*, and *cdtB* were determined by multiplex PCR as previously described.^16^ In addition, primers for *tcdC* were added as described by Persson et al.^17^ The PCR products were analyzed with the Agilent DNA 1000 assay and 2100 Bioanalyzer (Agilent Technologies, Waldbronn, Germany) to determine the length of the *tcdC* amplicon. Isolates with *tcdA, tcdB, cdtA, cdtB*, and a *tcdC* amplicon size of 144 base pairs were categorized as 027-like.

### Intervention and control arms

The intervention occurred at the level of ICU. During the baseline period, all ICUs followed their usual standard of care which consisted of healthcare workers following CDC contact precautions guidelines (gloves and gowns) for patients known to have antibiotic-resistant bacteria such as VRE, MRSA and *C. difficile* active infection.^18^ After the baseline period, ICUs were randomized, and during the study period, all healthcare workers (nurses, physicians, respiratory therapists, etc.) in the 10 ICUs assigned to the intervention arm were required to wear gloves and gowns for all patient contact and when entering any patient room.^18,19^ The 10 control ICUs followed their usual standard of care during the study period. Compliance with glove and gown use was measured by 30-minute direct observation periods on a random sample of rooms in both intervention units (any patient) and control units (patient on contact precautions). No hospitals performed active surveillance for *C. difficile*. All hospitals used contact precautions for the care of patients with clinical *C. difficile* infection. Twelve hospitals performed chlorhexidine bathing (five in the control arm and seven in the intervention arm).^20^

### Outcomes

For each eligible patient, acquisition was defined as having a baseline ICU surveillance peri-anal culture that was negative for toxigenic *C. difficile* with a subsequent discharge surveillance perianal culture within the same ICU admission that was positive for toxigenic *C. difficile* bacteria.

Results of clinical *C. difficile* testing results were not known to the study team and did not contribute to this definition.

Secondary analyses were conducted for factors associated with acquisition including ribotype 027-like *C. difficile* acquisition, colonization pressure, type of hospital ICU (medical, surgical or medical-surgical), hand hygiene, glove compliance, gown compliance, if patients were isolated for other reasons and month of intervention.

### Statistical analysis

The statistical analysis plan was written and sealed prior to the analysis. The analysis was based on the outcome (acquisition yes/no) for each person seen in the study ICUs at either the baseline period (when standard contact precautions were used in all ICUs) or the study period (when half of the ICUs employed universal contact precautions). The probability that each person was classified as having acquired an infection during their ICU stay is a function of the acquisition rate in that ICU at that period, and the number of days between admission specimen collection and discharge specimen collection which was approximately equal to the patient ICU length of stay. The rate of acquisition in an ICU at a given period was modeled as a multiplicative function of parameters related to period (baseline or study), contact precautions (whether that ICU was using universal or selective precautions during that period), and ICU (treated as a random effect). This corresponds to using a generalized linear mixed model for a binary outcome with a complementary log-log link, random effects for ICUs and the log of the number of days between swabs as an offset term. The model was fit by maximum likelihood estimation using SAS Proc GLIMMIX. The model resulted in estimates of the mean rate of acquisition during the baseline period, the mean rate during the study period in ICUs who performed selective precautions, the mean rate during the study period in ICUs who performed universal precautions, and the rate ratio due to the intervention.

For ease of interpretation, we also present a rate difference which is the difference in acquisition rates due to the intervention based on the model evaluated at the average ICU. Confidence intervals for the rate differences were calculated using the delta method based on the parameter estimates and standard errors from the multiplicative model that we fit.

The in-hospital colonization pressure was calculated for each patient as the proportion of other patients at their ICU who were positive for *C. difficile* during their stay (not including the patient themselves). Patients positive on admission were assumed to be positive their entire hospital stay. Patients who acquired *C. difficile* on discharge were assumed to acquire infection half-way through their stay.

#### Power calculations

Using assumptions based on the primary study, we assumed standard weighted averages of 0.12 and 0.13 in control and intervention ICUs with a baseline rate of 2% acquisition. We projected 80% power to detect an impact if the intervention reduced rates by 41%, performing two-sided 0.05-level test.

## RESULTS

Twenty ICUs participated in the study and none withdrew. Of the 26,749 patients enrolled in the study, 4,904 were excluded due to missing the admission or discharge culture result. A total of 21,845 patients had both admission and discharge perianal swabs worked up for toxigenic *C. difficile*, including 5912 patients during the baseline period and 15,933 patients during the study period. On admission toxigenic *C. difficile* was isolated from the rectal swab of 9.43% (2,060/21,845) patients. These were excluded from the analysis of hospital acquisition. During the study, compliance with obtaining perianal cultures at admission was 94.9%. Compliance with obtaining perianal cultures at discharge was 85.1%.

Compliance with wearing gloves in the intervention ICUs was 86.2% (2787/3234) and compliance with gowns was 85.1% (2750/3230). In the control group, 10.52% of patients were on contact precautions. In the control ICUs, for patients on contact precautions, compliance with wearing gloves was 84.11% (556/661) and compliance with gowns was 81.21% (536/660). Of patients who were found to be asymptomatically colonized with *C. difficile* on admission, in the baseline period 21% (176/822) were on contact precautions for various reasons. During the intervention period 18% (139/794) were on contact precautions for clinical reasons in control ICUs and 100% (902/902) were on contact precautions in intervention ICUs.

Figure 1 shows the baseline and study period rates of acquisition based on dividing the number of acquisitions by the days at risk in each ICU. Of the 19,785 patients not positive for *C. difficile* on admission, 6.6% (1,296/19,785) acquired *C. difficile*.

**Figure 1.**
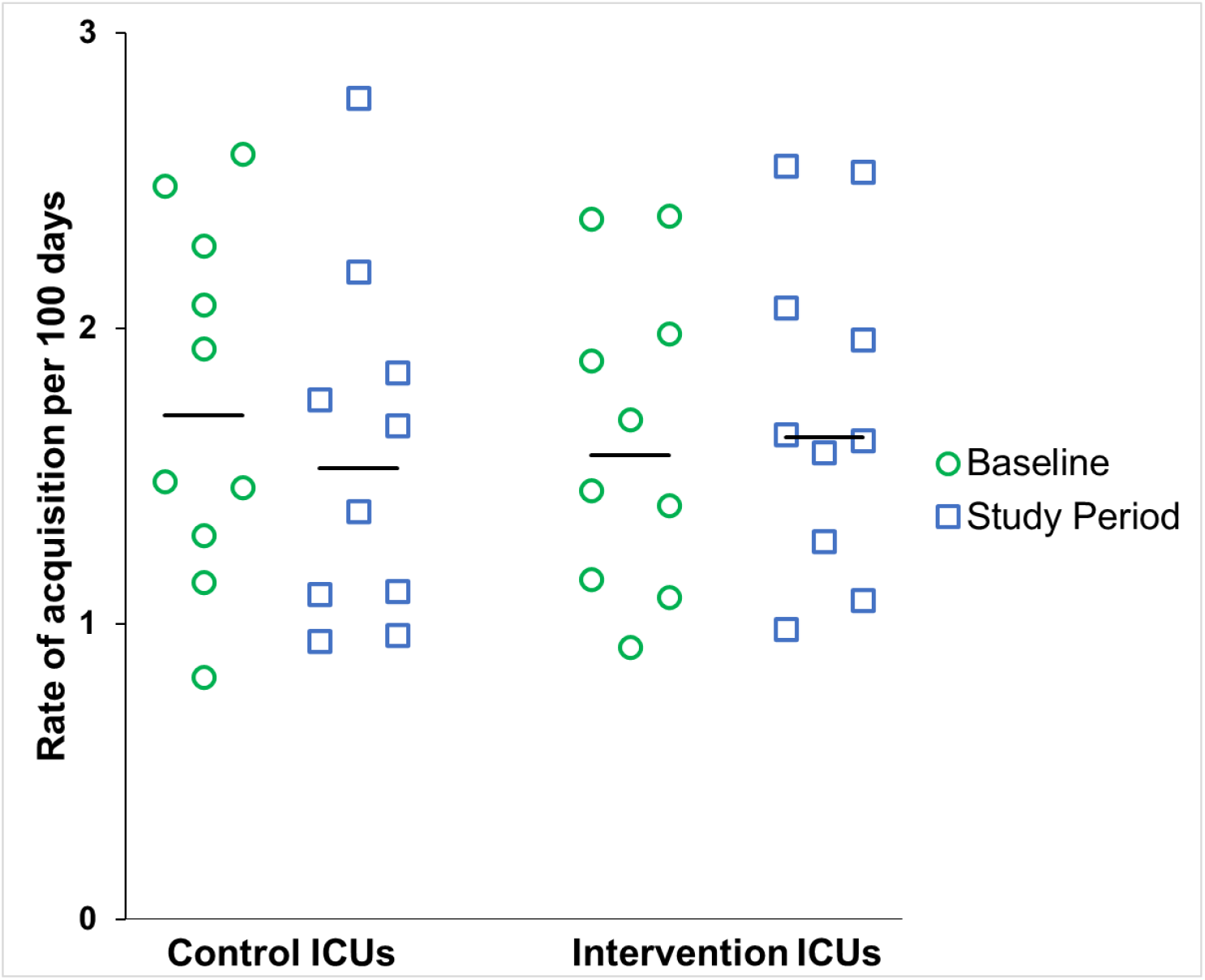
Rates of acquisition of toxigenic *C. difficile* per 100 days in the baseline and study periods for the control and intervention ICUs.

**Figure 2:**
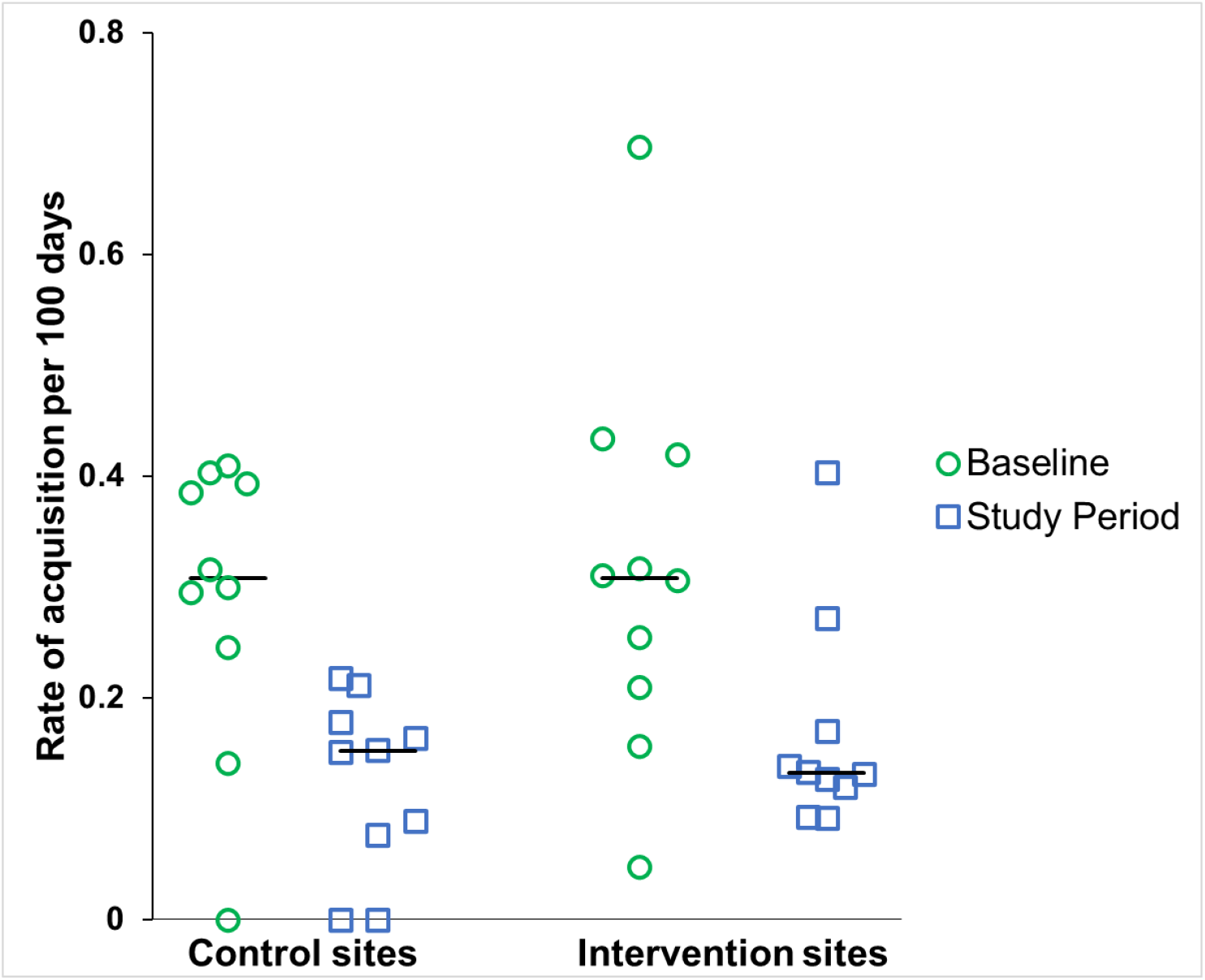
Rates of acquisition of 027-like *C. difficile* per 100 days in the baseline and study periods for the control and intervention ICUs.

Table 1 summarizes the primary outcome, rate of acquisition in intervention and control ICUs. During the study period, the rate of acquisition declined slightly in the control ICUs and increased slightly in the intervention ICUs, compared to the baseline period. Table 1 also provides the estimated rate difference due to the intervention based on the generalized linear mixed model. The estimated rate difference was -0.28 reflecting a lower rate in the control ICUs (p=0.091).

**Table 1.** Overall rate of acquisition in baseline and study periods, by intervention group.

|  | Baseline Period |  |  | Study Period |  |  | Rate Difference<sup>1</sup><br>(95% CI) | P-value<sup>1</sup> |
| --- | --- | --- | --- | --- | --- | --- | --- | --- |
|  | Acquisitions | Person-days | Rate per 100 person-days | Acquisitions | Person-days | Rate per 100 person-days |  |  |
| Control ICUs | 211 | 11,938 | 1.77 | 427 | 28,612 | 1.49 | -0.28 (-0.66, 0.10) | 0.091 |
| Intervention ICUs | 172 | 10,633 | 1.62 | 486 | 28,915 | 1.68 |  |  |

<sup>1</sup> Based on a generalized linear mixed model considering time between swabs and clustering within ICUs. Negative rate difference reflects lower rate in absence of intervention.

Secondary analyses were conducted to evaluate factors associated with acquisition. The colonization pressure experienced by each patient was defined as the proportion of other patients at the patient’s site that were presumed positive for *C. difficile* during the days that patient was in the ICU. Every 10% increase in colonization pressure was associated with 1.09 (1.02, 1.17) odds of increase in acquisition after adjusting for study period and calendar month, p=0.014)

Table 2 shows the relationship between other predictors and *C. difficile* acquisition for the entire study cohort. Time of year was also associated with *C. difficile* acquisition, with highest rates of acquisition observed in December and January, 2.71 and 2.79 per 100 patient days, p<0.001).

**Table 2.** Rates of acquisition in subgroups defined by site characteristics and month of study.

| Predictor | Subgroup | Number of Patients | Acquisitions | Person-days at risk | Rate (per 100 person-days) | P-value |
| --- | --- | --- | --- | --- | --- | --- |
| Type of ICU | MICU | 8757 | 595 | 38525 | 1.54 | 0.15 |
|  | MICU/SICU | 4679 | 300 | 15695 | 1.91 |  |
|  | SICU | 6349 | 401 | 25879 | 1.55 |  |
| Hand Hygiene at entry | 55% or greater | 10939 | 747 | 41821 | 1.79 | 0.15 |
|  | <55% | 8846 | 549 | 38277 | 1.43 |  |
| Glove compliance | 75% or greater | 16889 | 1127 | 67756 | 1.66 | 0.21 |
|  | <75% | 2896 | 169 | 12342 | 1.37 |  |
| Gown Compliance | 75% or greater | 16028 | 1070 | 62120 | 1.72 | 0.03 |
|  | <75% | 3757 | 226 | 17978 | 1.26 |  |
| Patient Isolated* | No | 11007 | 693 | 43060 | 1.61 | 0.99 |
|  | Yes | 8725 | 599 | 36847 | 1.63 |  |
| Year/month | August 2011 | 67 | 3 | 291 | 1.03 | <0.001 |
|  | September 2011 | 1020 | 63 | 4682 | 1.35 |  |
|  | October 2011 | 1361 | 58 | 5674 | 1.02 |  |
|  | November 2011 | 1340 | 105 | 5932 | 1.77 |  |
|  | December 2011 | 1349 | 147 | 5422 | 2.71 |  |
|  | January 2012 | 1516 | 170 | 6104 | 2.79 |  |
|  | February 2012 | 1460 | 133 | 7190 | 1.85 |  |
|  | March 2012 | 1718 | 98 | 6764 | 1.45 |  |
|  | April 2012 | 1601 | 90 | 6404 | 1.41 |  |
|  | May 2012 | 1648 | 72 | 6752 | 1.07 |  |
|  | June 2012 | 1717 | 97 | 6764 | 1.43 |  |
|  | July 2012 | 1646 | 116 | 6427 | 1.81 |  |
|  | August 2012 | 1738 | 64 | 6701 | 0.96 |  |
|  | September 2012 | 1513 | 79 | 4874 | 1.62 |  |
|  | October 2012 | 91 | 1 | 120 | 0.84 |  |

\*reasons for isolation included MRSA, VRE, *C. difficile* infection. 53 patients were missing isolation status.

We evaluated the impact of universal gowns and gloves on 027-like *C. difficile* and found there was no significant difference between intervention and control ICUs. Rates of 027-like *C. difficile* in control ICUs were 0.28 per 100 patient days during baseline and 0.13 per 100 patient days during the study period vs. intervention ICUs were 0.30 per 100 patient days during baseline and 0.16 per 100 patient days during the study period, rate difference -0.03 (95% CI - 0.11, 0.05), p-value 0.35, generalized linear mixed effects model.

## DISCUSSION

In a cluster randomized trial of 20 ICUs we found no evidence that universal gown and glove use had an impact on acquisition rates of toxigenic *C. difficile* or ribotype 027-like *C. difficile*. Other factors associated with *C. difficile* acquisition included *C. difficile* colonization pressure in the ICU, self-reported high compliance with gown and glove use and time of year.

The optimal approach to preventing CDI has been debated.^7,21,22^ Most current infection prevention efforts focus on patients with CDI and not asymptomatic carriers.^22^ However, asymptomatic patients can shed spores into the environment^21^ and a before-after study in one hospital found lower rates of CDI with active surveillance and isolation of patients who were asymptomatic.^7^ In our 20 hospital study, following a baseline period, 10 ICUs were randomized to universal glove and gown use and 10 served as controls. We observed no effect in the comparison of ICUs randomized to intervention vs. standard care. It is notable that 18% of patients who were colonized with *C. difficile* at admission were on contact precautions for clinical indications in standard care ICUs vs. 100% of patients who were on contact precautions in intervention ICUs. Notably, there was no difference in outcome despite this difference. These results suggest that active surveillance and isolation of *C. difficile* may not prevent *C. difficile* beyond current standard practice of isolating patients who have active diarrhea. Likewise, the intervention had no impact on *C. difficile* acquisition of strains characteristic of ribotype 027. While this argues against a potential benefit of identifying and isolating asymptomatic *C. difficile* colonized patients, there may be other measures associated with active detection of colonized patients, in particular - the use of sporicidal agents for environmental disinfection, that could not be accounted for in this study.

We found interesting associations on secondary analyses. These include an apparently contradictory finding of lower *C. difficile* acquisition in ICUs that reported lower levels of compliance with gown and gloves. We do not believe that worse use of gowns and gloves prevents *C. difficile* acquisition but that, perhaps those ICUs who were more rigorous about collecting true compliance had lower rates of *C. difficile* acquisition. Other associations included the observation that ICUs with a greater proportion of patients with *C. difficile* colonization had greater rates of *C. difficile* acquisition. This phenomenon of colonization pressure is well documented for multiple organisms, including *C. difficile*.^23,24^ However, it is unclear whether this represents true patient-to-patient transmission risk or whether the higher burden of asymptomatic *C. difficile* colonization is a marker for other risk factors for CDI such as high antimicrobial use. We also observed variation in acquisition of *C. difficile* by time of year with highest rates of acquisition in early winter. Seasonal variability of CDI has been observed in the past and, in part, was explained by increased influenza and viral illness leading to increased antibiotic use.^25^ The 2011-2012 season had increased rates of influenza-like illness starting December of 2011.^26^

The impact of universal gown and glove use has been explored in the BUGG study. The primary study found no overall difference in rates of MRSA or VRE acquisition, but an a priori defined secondary analysis identified an absolute reduction in MRSA (4.03% absolute risk reduction, p=0.046) and no impact on VRE acquisition. Gram-negative bacteria were likewise not statistically significantly impacted by universal gown and glove, 6.48% absolute risk increase, p=0.09).^12^ Taken as a whole, it appears that universal gown and glove use, may have some impact on MRSA acquisition, which is a skin organism and potentially more responsive to contact precautions, and no impact on organisms primarily found in the GI tract, such as VRE, gram-negative rods and *C. difficile*.

Although our study had strengths coming from the 20-ICU randomized controlled trial design, there were limitations as part of the limited identifying information of the trial including that we did not have individual patient level data for CDI or antibiotic use. Likewise, we used peri-anal swabs that are less sensitive than stool or rectal swabs. We do not think any of these limitations would be differential between groups.

In conclusion, in a cluster randomized trial of universal glove and gown use, we found no benefit to the isolation of patients asymptomatically colonized with *C. difficile* compared to standard practice of isolating patients with CDI. This supports the current practice of patient isolation for symptomatic CDI.

## Data Availability

Limited data produced in the present study are available upon reasonable request to the authors

## Disclaimer

Statements in the manuscript do not necessarily represent the official views of, or imply endorsement by, AHRQ or HHS.

